# Prognostic implications of intraventricular conduction disorders for sudden cardiac death in coronary artery disease

**DOI:** 10.1101/2023.11.01.23297945

**Authors:** Roope Lahti, Jani Rankinen, Minna Järvensivu-Koivunen, Juho Tynkkynen, Markku Eskola, Kjell Nikus, Jussi Hernesniemi

**Affiliations:** Faculty of Medicine and Health Technology and Finnish Cardiovascular Research Center Tampere, Tampere University, Tampere, Finland; Heart Center, Department of Cardiology, Kanta-Häme Central Hospital, Hämeenlinna, Finland; Heart Hospital Department of Cardiology, Tampere University Hospital, Tampere, Finland; Department of Radiology, Tampere University Hospital, Tampere, Finland

## Abstract

**Backround and aims:** Prolonged QRS duration (≥ 110 ms) and coronary artery disease are risk factors for sudden cardiac death (SCD). We explored the SCD risk associated with intraventricular conduction disorders – a prolonged QRS duration of 110–119 ms, right bundle branch block (RBBB), left bundle branch block (LBBB), and a nonspecific intraventricular conduction delay (NIVCD) – in patients with acute coronary syndrome (ACS).

**Methods:** This is a retrospective study of 9,700 consecutive, invasively treated ACS patients with ECGs available for analysis (2007–2018). SCD definition was based on an in-depth review of written medical records and death certificates describing the circumstances leading to the events. Endpoint data were available until December 31, 2021 (no losses to follow-up). The risk associated with conduction disorders was analyzed by calculating subdistribution hazard estimates (deaths due to other causes being considered competing events).

**Results:** The median follow-up time was 6.8 years (IQR 4.0–10.2), during which 3,420 (35.3%) patients died. SCDs were overrepresented as a cause of death among patients with NIVCD (16.4%) or with a prolonged QRS duration (15.3%) when compared to patients with LBBB (5.3%), RBBB (7.1%), or with a normal QRS duration (10.5%). In an analysis adjusted for age, sex, and cardiac comorbities, NIVCD and a prolonged QRS were significant predictors of SCD (HR 3.00, 2.06– 4.35, P < 0.001; and HR 1.80, 1.37–2.35, P < 0.001, respectively). After adjusting the analysis with left ventricular ejection fraction, NIVCD and a prolonged QRS duration remained as significant risk factors for SCD. LBBB and RBBB did not predict SCD.

**Conclusion:** The incidence of SCD is significantly higher in patients with NIVCD and a prolonged QRS duration. Approximately 23% of all SCDs occur among these patients.

## Introduction

Sudden cardiac death (SCD) is a global problem and usually a manifestation of acute coronary syndrome (ACS) events caused by coronary artery disease (CAD) (1). Even after optimal revascularization and secondary prevention, CAD patients are at risk of SCD because of recurring ischemic events and incidental ventricular arrhythmias (2). For example, depending on the estimate, 12%–35% of deaths after an ACS event are SCDs, and the annual incidence is roughly 0.4%–1.0% (3–5). For comparison, the annual incidence of SCD in the general population is only 0.03%–0.10% (5–8). Due to their high risk of SCD, ACS patients are an optimal population for the search of new SCD risk markers and improvements in risk stratification for the primary prevention of SCD by an implantable cardioverter-defibrillator (ICD). Currently, the main criterion for primary prevention with an ICD is a left ventricular ejection fraction (LVEF) of under 35% despite optimal heart failure medication and revascularization (9). However, only a small fraction of SCDs can be prevented with these criteria, and, therefore, additional risk markers are needed to improve the risk stratification (4,10–12).

A prolonged QRS duration (≥ 110 ms) in an electrocardiogram (ECG) has been linked to an increased SCD risk in general population samples (13–15). The relationhip between conduction disorders (right bundle branch block [RBBB], left bundle branch block [LBBB], and a non-specific intraventricular conduction delay [NIVCD]) and SCD is less studied, and the results are controversial (16–18). There is initial proof that NIVCD patients suffer more arrhythmic deaths (13,19).

The purpose of this study was to investigate the implications of different intraventricular conduction disorders (prolonged QRS ≥ 110 ms, RBBB, LBBB, NIVCD) for SCD risk stratification in a large contemporary cohort of patients treated for ACS.

## Methods

### Study population

The study population consist of the retrospective data of consecutive patients undergoing invasive evaluation for ACS at Tays Heart Hospital between January 1, 2007, and December 31, 2018. Tays Heart Hospital is the sole provider of acute invasive cardiologic care for patients suffering from ACS in the Tampere Region with a population of roughly 0.5 million. The follow-up for mortality lasted from the first hospitalization (beginning from January 1, 2007) until December 31, 2021. During the study period (2007–2018), a total of 11,352 angiographies were performed for acute coronary syndrome on 10,314 patients. After excluding patients with no recorded ECG within the predefined time frame (1 week before or 2 months after angiography, n = 462), those with a ventricular-paced ECG (n = 149), or those with ventricular pre-excitation (n = 3), and accepting only the first ACS as the index event for each patient, 9,700 patients were followed for sudden cardiac death. The majority (n = 9,329, 96.0%) of the ECG recordings were taken on the same day as or within one week after the angiography (same day, n = 1,120, 11.5%; 1–7 days after the angiography, n = 8,209, 84.5%; 8 days–2 months after the angiography, n = 372, 3.8%). Only twelve (0.1%) ECGs were taken before the day of the angiography (1–3 days).

Patients not diagnosed invasively by means of coronary angiography were not included in the study (less than 10% of all ACS patients in the study center) (20). This exclusion was made because the diagnosis of ACS in patients who have not undergone an invasive evaluation is uncertain and the treatment strategy was usually based on a poor overall prognosis (20,21). This study is part of the retrospective MADDEC (Mass Data in Detection and Prevention of Serious Adverse Events in Cardiovascular Disease) study, which aims to utilize mass data for the prediction and prevention of serious cardiovascular adverse events (22).

### Baseline phenotype data collection

The baseline data were extracted from the MADDEC database, which retrospectively combines data collected from the 1990s onwards from different electronic databases used in specialized health care to create a comprehensive study registry focusing on patients treated at Tays Heart Hospital. The database combines electronic health records (EHR) with data from the prospectively collected and actively maintained KARDIO database (data collection performed by physicians and nurses during the treatment of patients) and with data gathered retrospectively by physicians using a full-disclosure review of written health care records. ACS and its subtypes were defined by European Society of Cardiology (ESC) and American College of Cardiology (ACC) criteria (23–25).

### Main exposure variables

The main exposure variables were a QRS duration of 110–119 ms, LBBB, RBBB (with or without left anterior/posterior fascicular block), and NIVCD. LBBB was defined as a QRS duration of ≥ 140 ms for men and ≥ 130 ms for women, along with QRS notching or slurring in ≥ 2 contiguous leads (26). RBBB required a QRS duration ≥ 120 ms, as well as rsr′, rsR′, rSR′, or qR in leads V1 or V2, and wide S waves in leads I, V5, and V6 (27). Non-specific IVCD was defined as QRS ≥120 ms without typical features of LBBB or RBBB. Figure 1 presents examples of three NIVCD demonstrations.

**Figure 1:**
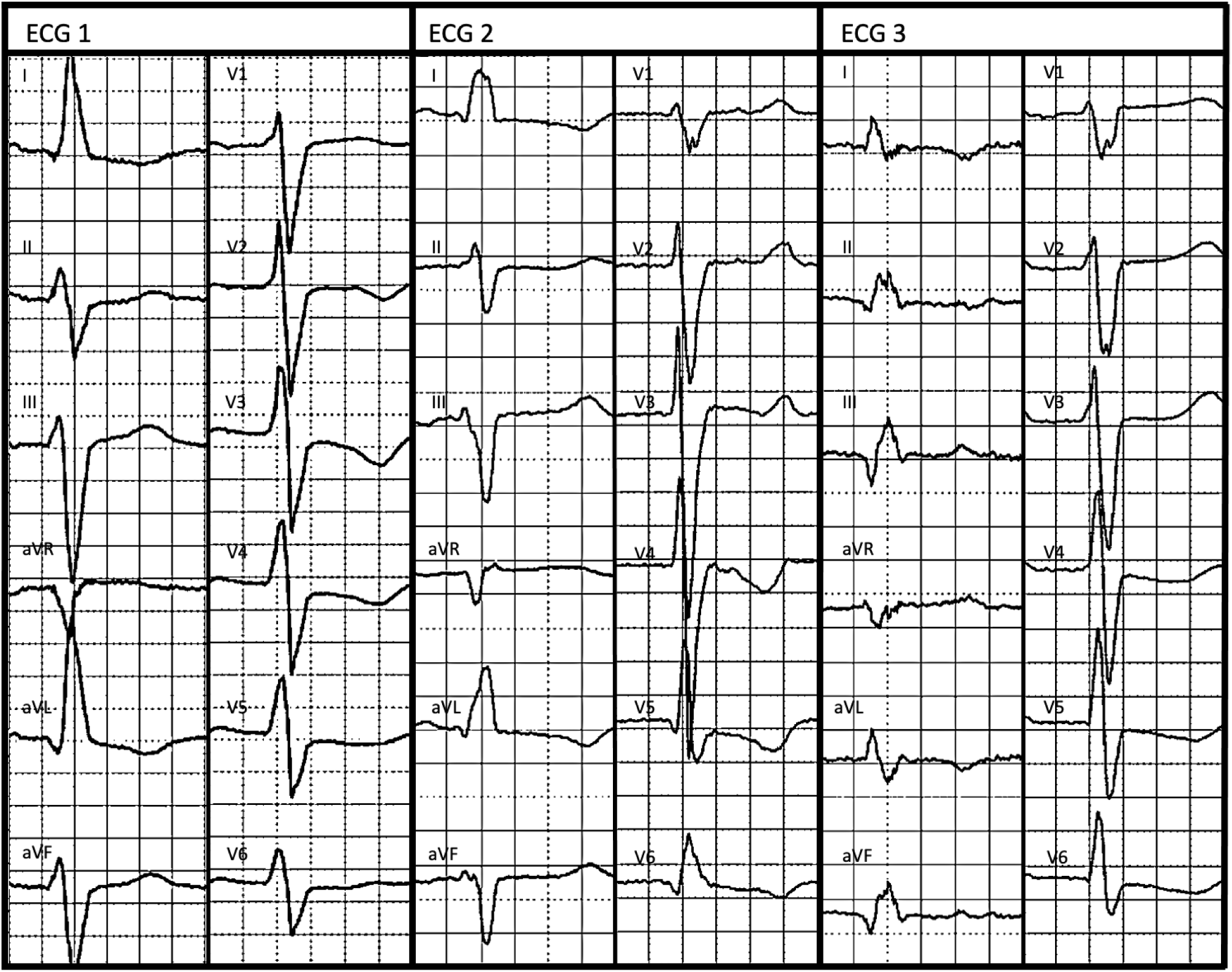
NIVCD ECG demonstrations.

**Figure 2:**
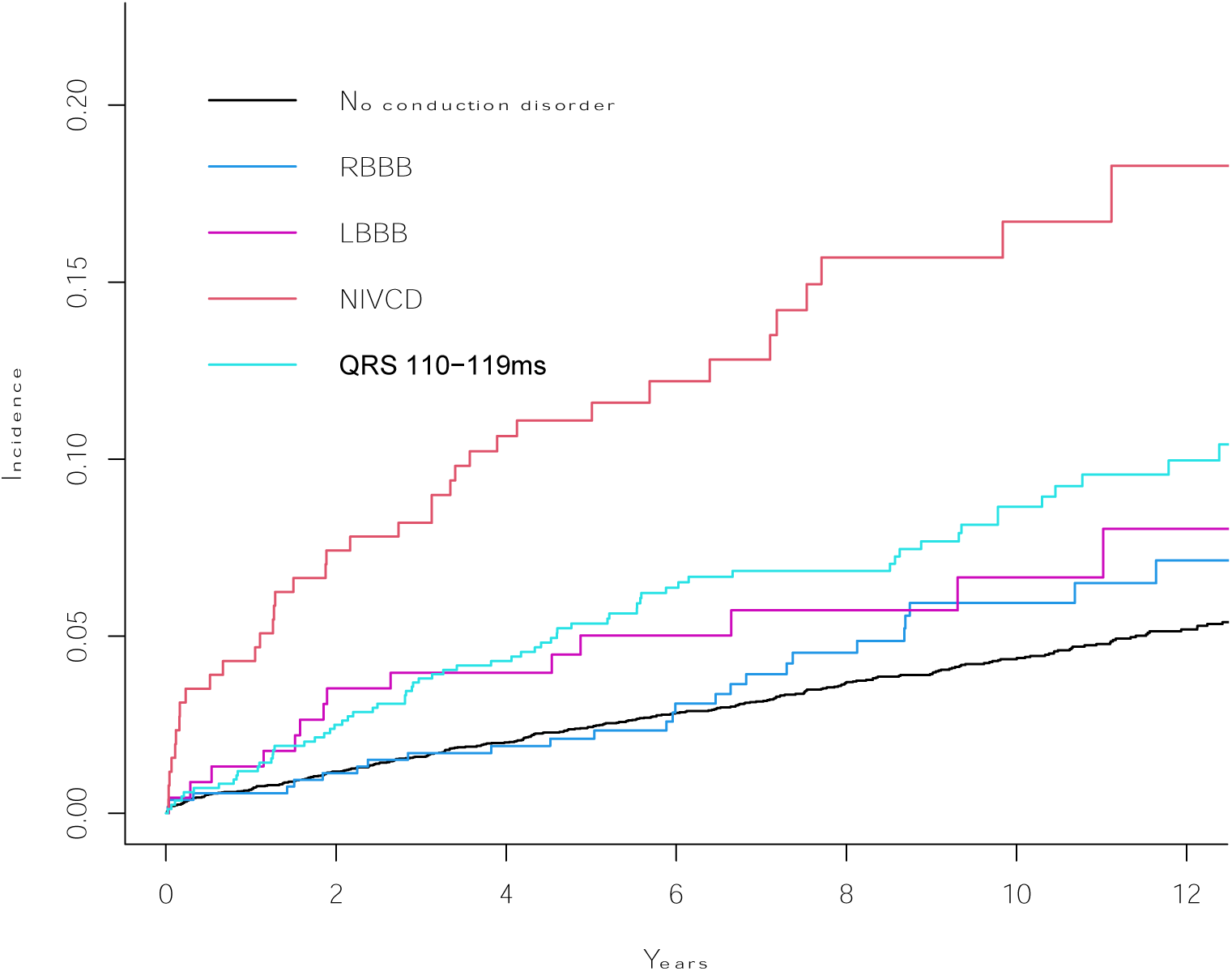
Risk of SCD by conduction disorder groups in ACS patients.

**Figure 3:**
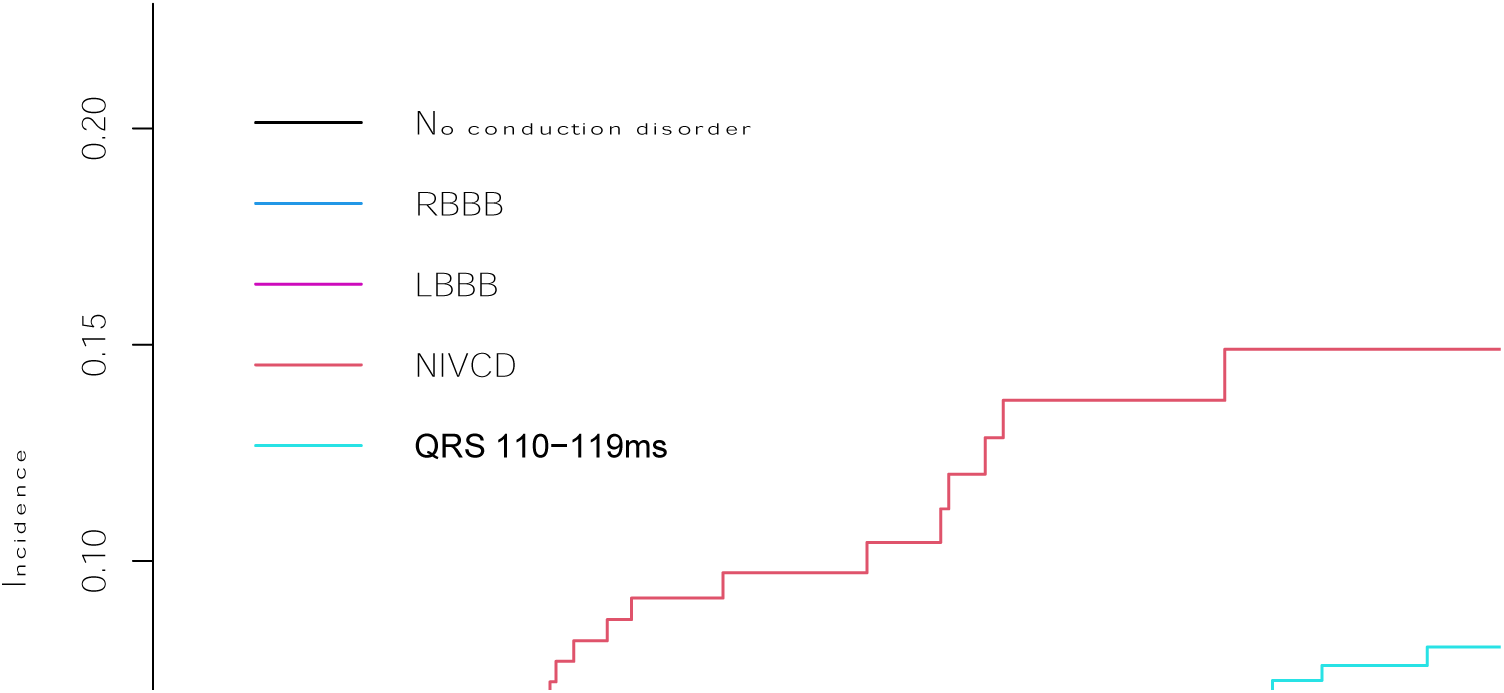
Risk of SCD by conduction disorder groups in patients without ICD.

**Figure 4:**
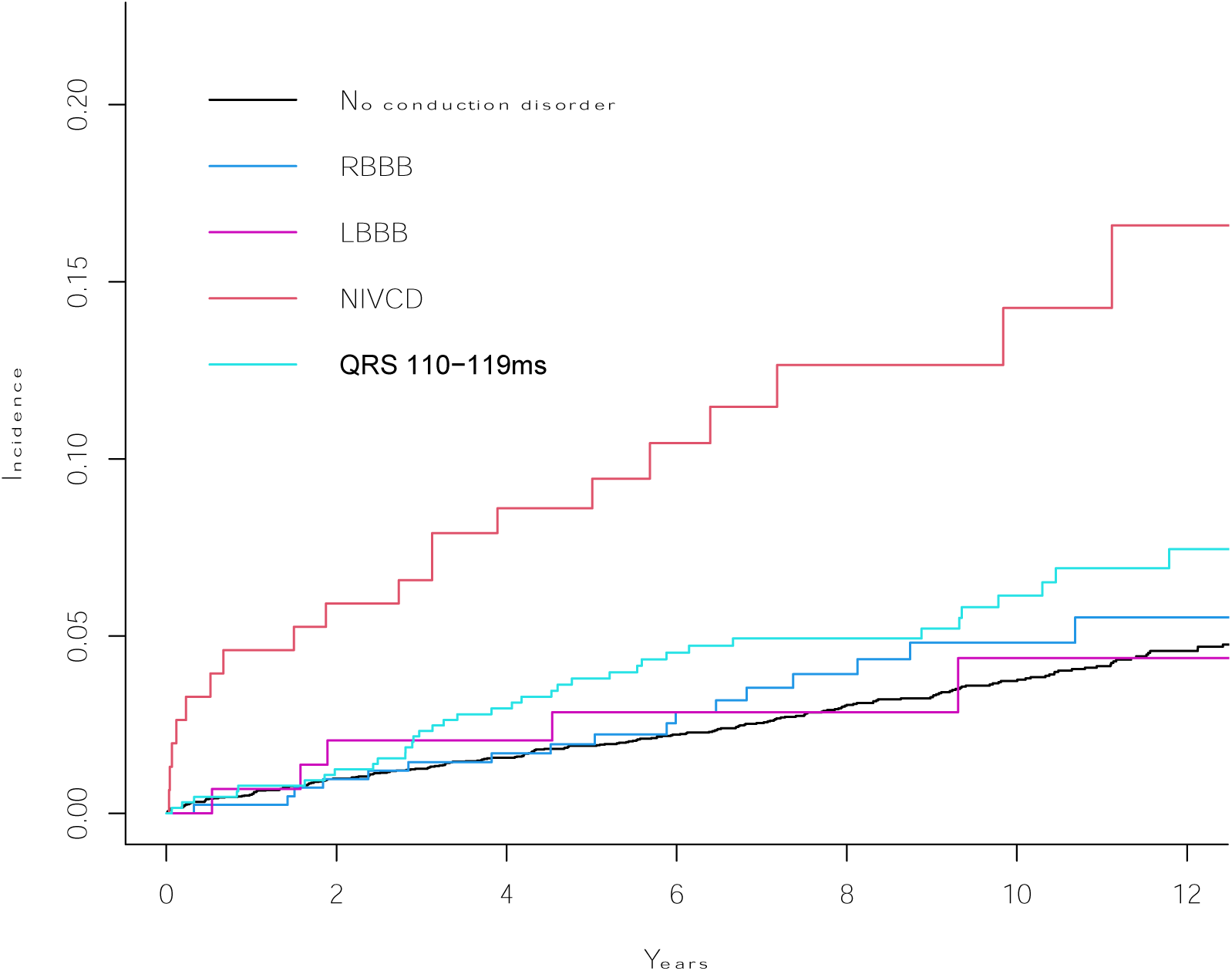
Risk of SCD by conduction disorder groups in patients with LVEF>35.

All ECGs were first classified with 12SL/GE Marquette software. All ECGs with a statement of RBBB, LBBB, NIVCD, incomplete (i)RBBB, iLBBB, left anterior fascicular block (LAFB), left posterior fascicular block (LPFB), and QRS ≥120 ms were manually classified by researchers (R.L. and J.R.) and categorized (QRS duration also evaluated manually). In the case of disagreement, a third cardiologist was consulted (K.N.). Patients were classified into five categories: normal QRS (QRS < 110 ms), prolonged QRS 110–119 ms, RBBB, LBBB, and NIVCD. We manually evaluated whether the conduction disorders (RBBB, LBBB, and NIVCD) were old, new, or presumabely new (no ECG available) from ECGs taken before discharge. The classification of conduction disorders by the 12SL Marquette system compared to a clinical evaluation is presented in Supplementary Table 1.

### Follow-up and endpoint definitions

The follow-up for each patient lasted from the index event to death or until the end of the year 2021 (December 31, 2021). The main endpoint was a true SCD or an equivalent event occurring after discharge. Deaths due to cardiac or non-cardiac causes were also followed. The definitions of SCD (and sudden cardiac arrest [SCA]) were based on the AHA/ACC/HRS and ESC guidelines (28,29). SCD is defined as a sudden and unexcepted death occurring within one hour of symptom onset, or within 24 hours of the individual having been observed asymptomatic and due to a cardiac cause. An SCD-equivalent event was defined as a similar event that would have resulted in a true SCD (symptom onset < 1 h) but was aborted by a successful resuscitation. Additionally, we also included all fast ventricular tachycardias adequately treated by an ICD device and resulting in a hemodynamic collapse or ventricular fibrillation (VF) episodes as SCD-equivalent events. SCA (a secondary endpoint) was defined as hemodynamic collapse due to a sudden cessation of cardiac activity because of a presumably cardiac cause (28,29). The determination of SCD/SCA was based on an in-depth review of written hospital medical records and death certificates detailing the circumstances leading to the events by two investigators, M.J-K and J.H. (4). If the written medical records or death certificate did not indicate the death being sudden or cited a non-cardiac cause (such as trauma, drowning, respiratory failure or asphyxia, electrocution, drug overdose, or other noncardiac cause), the death was not recorded as an SCD.

SCD cases that were not unexpected were recorded as (cardiac) deaths or sudden cardiac arrest events, such as in the case of prolonged cardiac symptoms or a vague event description, poor functional capacity or dementia, a poor clinical condition, or if the patient was in palliative care.

General causes of death data were retrieved from Statistics Finland in International Classification of Diseases, Tenth Revision (ICD–10) format. The causes of death register by Statistics Finland provides 100% coverage of all deaths of citizens and permanent residents of Finland. Consequently, there were no losses to follow-up. The cause of death is reported to the registry according to the ICD code of the primary or immediate cause of death, which is extremely precise in Finland. Patients whose cause of death was coded as I20–I52 were further classified as having died of a cardiac cause and as SCD if the above-mentioned criteria were fulfilled.

### Statistical analysis

Comparisons between the patient groups were performed with normal Chi-squared testing for categorical variables, with ANOVA for normally distributed continuous variables, and with the Kruskall–Wallis or Mann–Whitney U test for non-normally distributed continuous variables. The cumulative incidence of SCDs and the prognostic value of conduction disorders during the entire follow-up were modeled using unadjusted and adjusted (continuous variables entered into the models without categorization) Fine–Gray subdistribution hazard models, which account for competing risk due to other causes of death (30), and with traditional Cox regression, which does not take competing events into account. The analyses were performed with SPSS (version 27, IBM) and R software (version 4.1.3; packages survival and cmprisk).

## Results

### General characteristics of the study population and events during follow-up

The mean age of the entire population at baseline was 68.2 (11.8) years, and 67.3% (n = 6,532) of the patients were men. The median follow-up time was 6.8 years (IQR 4.0–10.2), during which 3,420 (35.3%) patients died. Approximately half (51.4%, n = 1,757) of the patients died of cardiac causes. During follow-up a total of 578 SCA events were recorded (occurring in 6.0% of all patients), 65.2% (n = 377) of which were classifiable as true, fatal SCDs. The majority (96.8 %, n = 365/377) of the true SCDs occurred among patients with no ICD device. In addition, there were 461 SCD or SCD-equivalent events (79.8% of all SCAs), and in 4.3% (n = 20/461) of these cases, the patient was successfully resuscitated, while 14.1% (n = 65/461) had fast ventricular tachycardias (VT) or ventricular fibrillation (VF) that were treated succesfully with an ICD. Out of those who were successfully resuscitated or received adequate ICD therapy, 95.3% (n = 81/85) survived for more than 30 days.

### Intraventricular conduction disorders in patients with ACS

Of all ACS patients, 8.7% (n = 840) had a QRS duration of 110–119 ms, 5.5% (n = 310) had RBBB, 2.3% (n = 227) had LBBB, and 2.6% (n = 256) had NIVCD in their ECG (Table 1). Approximately half (49.0%, n = 496/1013) of the conduction blocks were new (n = 212) or presumably new (n = 284). In general, patients with a prolonged QRS duration of 110–119 ms, RBBB, LBBB, or NIVCD were older and had a higher prevalence of comorbidities at baseline, when compared to patients with no observable conduction disorders (Table 1). The classification of NIVCDs by the 12SL Marquette system compared to a clinical evaluation is presented in Supplementary Tables 1 and 2.

**Table 1.**
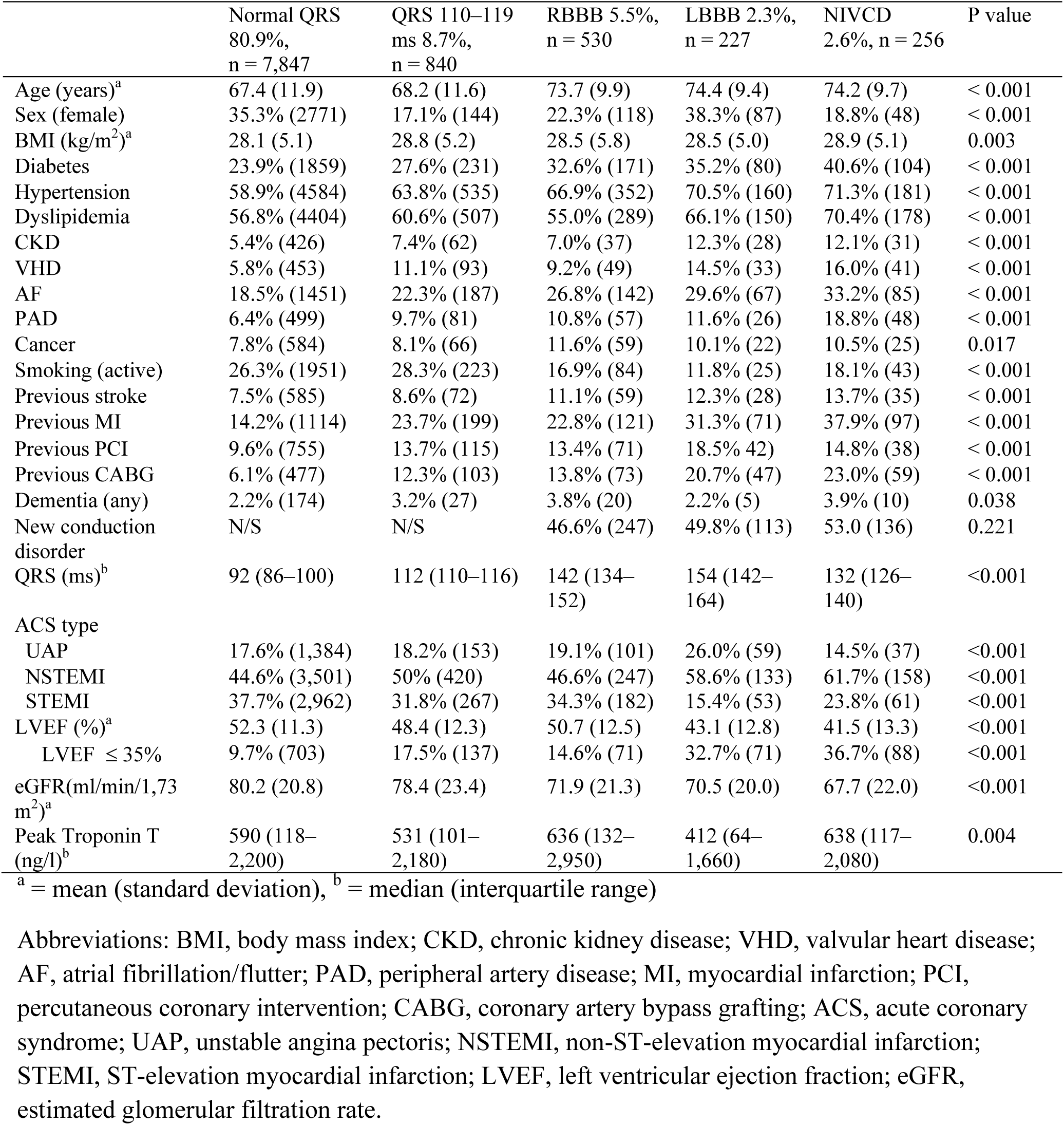
General characteristics of the study population.

### Intraventricular conduction disorders and SCD events during follow-up

Patients with intraventricular conduction disorders, who were also older, had a significantly higher (p < 0.001) all-cause mortality (ranging from 42.7% for prolonged QRS duration to 68.8% for NIVCD), when compared to patients with a normal QRS complex (31.8%) (Table 2). True fatal SCDs were overrepresented as a cause of death among patients with NIVCD (16.4%) or a prolonged QRS duration (15.3%), when compared to patients with LBBB (7.1%), RBBB (7.6%), or with a normal QRS duration (10.6%) (Table 2). Out of all SCDs, 32% occurred in patients with conduction disorders (LBBB, RBBB, NIVCD, or prolonged QRS duration of 110–119 ms) and 23% in patients with NIVCD or a prolonged QRS duration (110–119 ms). The corresponding percentages were 28% and 21%, respectively, when focusing only on patients with no ICDs.

**Table 2.** Events during follow-up stratified by intraventricular conduction block.

|  | QRS <110ms<br>n = 7,847 | QRS<br>110–119 ms<br>n = 840 | RBBB<br>n = 530 | LBBB<br>n = 227 | NIVCD<br>n = 256 |
| --- | --- | --- | --- | --- | --- |
| SCD or equivalent event,<br>% of all subjects (n) | 4.0% (313) | 8.3% (70) | 5.1% (27) | 6.2% (14) | 14.5% (37) |
| Fatal SCD, % of all deaths<br>(n) | 10.6% (264) | 15.3% (55) | 7.6% (21) | 7.1% (8) | 16.4% (29) |
| Resuscitated SCD, % of<br>SCD/equivalent events (n) | 5.1% (16) | 2.9 (2) | 0 | 14.3% (2) | 0 |
| Adequate ICD therapy*, %<br>of ICD patients (n) | 10.5% (33) | 18.6% (13) | 22.2% (6) | 28.6% (4) | 24.3% (9) |
| SCD or equivalent event<br>in patients with no ICD (n) | 3.5% (272) | 6.5% (54) | 3.6% (19) | 3.1% (7) | 10.8% (27) |
| Any SCA event | 5.7% (445) | 11.3% (95) | 9.2% (49) | 11.5% (26) | 20.3% (52) |
| Total deaths | 31.8% (2495) | 42.7% (359) | 52.5% (278) | 49.3% (112) | 68.8% (176) |
| ICD devices overall | 2.3% (184) | 5.4% (45) | 4.9% (26) | 13.2% (30) | 12.9% (33) |
| Previous ICD | 0.1% (7) | 0.5% (4) | 0.2% (1) | 0.9% (2) | 2.3% (6) |
| Incident ICD | 2.2% (174) | 4.9% (41) | 4.7% (25) | 12.3% (28) | 10.5% (27) |

\*Adequate ICD therapy for fast ventricular tachycardia leading to hemodynamic collapse, or for VF. In one patient, an adequate ICD shock occurred simultaneously with true SCD.

ICD implantation was more prevalent in patients with NIVCD and LBBB than other patient groups (Table 2). Overall ICD therapies accounted for 17.3% of events classified as SCDs or equivalent events among patients with conduction disorders, whereas in patients with a QRS duration of < 110 ms, ICD therapies accounted for 10.5% of the events (p = 0.01)(Table 2).

In the Fine-Gray subdistribution hazard (SDH) model, NIVCD and prolonged QRS, but not RBBB or LBBB, were associated with SCD in the non-adjusted analysis (Table 3). NIVCD and a prolonged QRS duration of 110–119 ms remained as significant predictors of SCD when adjusting for age, sex, and cardiac comorbidities (NIVCD: HR 3.00, 95% CI 2.06–4.35, P < 0.001; prolonged QRS: HR 1.80, 1.37–2.35, P <0.001), as well as age, sex, and LVEF (HR 2.60, 1.78–3.81, P <0.001, and HR 1.76, 1.35–2.30, P <0.001, respectively). The same applied when the analysis was restricted to patients with a LVEF of > 35% and adjusted for age and sex (HR 3.53, 2.18–2.34, P <0.001, and HR 1.66, 1.18–2.34, P <0.001, respectively) (Table 3). The traditional Cox regression analysis yielded similar results (Table 4).

**Table 3.**
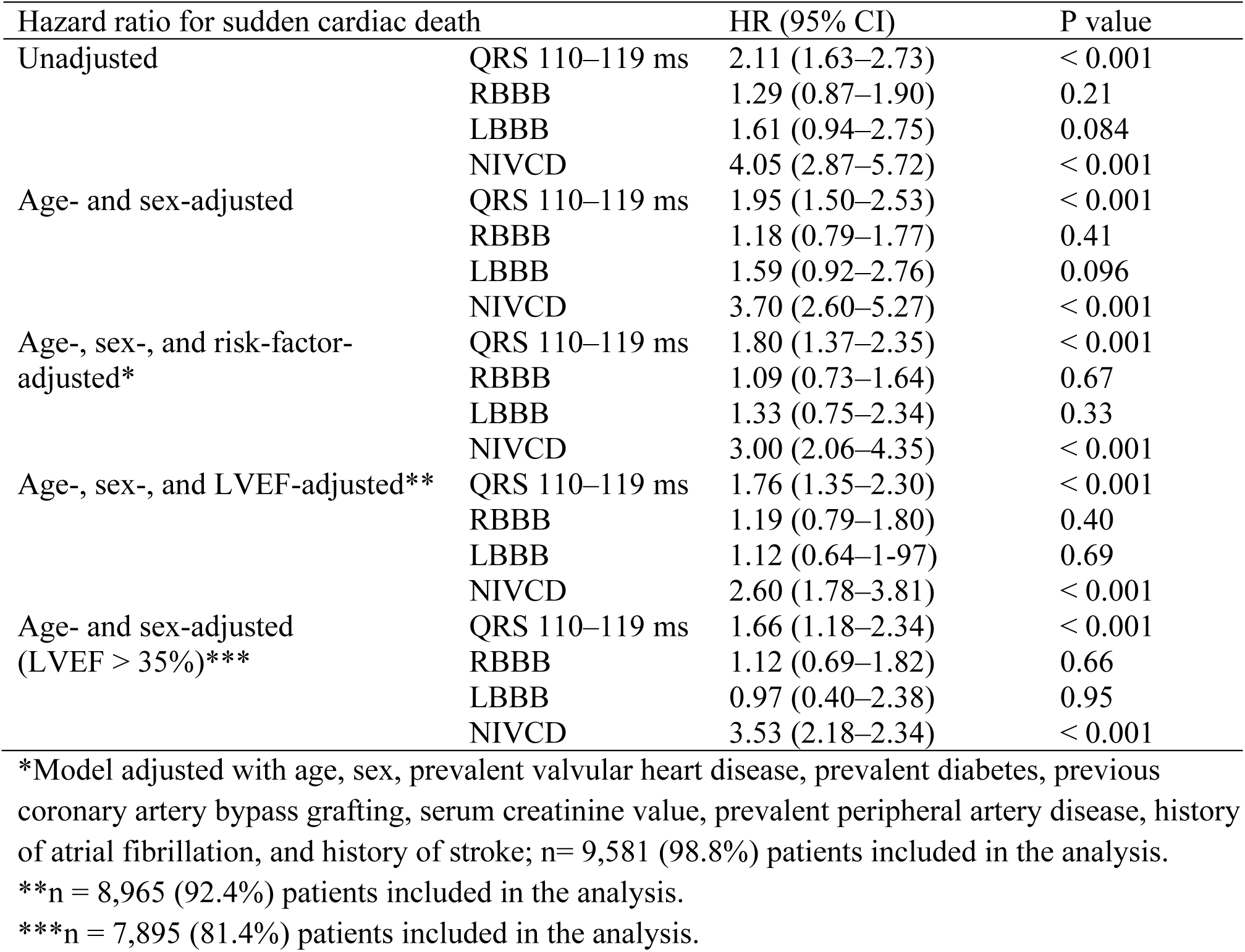
The association between intraventricular conduction disorders and the incidence of sudden cardiac death among patients treated for acute coronary syndrome. Hazard ratio estimates (HRs) correspond to subdistribution hazard estimates analyzed by Fine-Gray subdistribution hazard modeling, accounting for competing events due to deaths due to other causes.

| Hazard ratio for sudden cardiac death |  | HR (95% CI) | P value |
| --- | --- | --- | --- |
| Unadjusted | QRS 110–119 ms | 2.11 (1.63–2.73) | < 0.001 |
|  | RBBB | 1.29 (0.87–1.90) | 0.21 |
|  | LBBB | 1.61 (0.94–2.75) | 0.084 |
|  | NIVCD | 4.05 (2.87–5.72) | < 0.001 |
| Age- and sex-adjusted | QRS 110–119 ms | 1.95 (1.50–2.53) | < 0.001 |
|  | RBBB | 1.18 (0.79–1.77) | 0.41 |
|  | LBBB | 1.59 (0.92–2.76) | 0.096 |
|  | NIVCD | 3.70 (2.60–5.27) | < 0.001 |
| Age-, sex-, and risk-factor-adjusted* | QRS 110–119 ms | 1.80 (1.37–2.35) | < 0.001 |
|  | RBBB | 1.09 (0.73–1.64) | 0.67 |
|  | LBBB | 1.33 (0.75–2.34) | 0.33 |
|  | NIVCD | 3.00 (2.06–4.35) | < 0.001 |
| Age-, sex-, and LVEF-adjusted** | QRS 110–119 ms | 1.76 (1.35–2.30) | < 0.001 |
|  | RBBB | 1.19 (0.79–1.80) | 0.40 |
|  | LBBB | 1.12 (0.64–1.97) | 0.69 |
|  | NIVCD | 2.60 (1.78–3.81) | < 0.001 |
| Age- and sex-adjusted (LVEF > 35%)* | QRS 110–119 ms | 1.66 (1.18–2.34) | < 0.001 |
|  | RBBB | 1.12 (0.69–1.82) | 0.66 |
|  | LBBB | 0.97 (0.40–2.38) | 0.95 |
|  | NIVCD | 3.53 (2.18–2.34) | < 0.001 |

\*Model adjusted with age, sex, prevalent valvular heart disease, prevalent diabetes, previous coronary artery bypass grafting, serum creatinine value, prevalent peripheral artery disease, history of atrial fibrillation, and history of stroke; n = 9,581 (98.8%) patients included in the analysis.
\*\*n = 8,965 (92.4%) patients included in the analysis.
\*\*\*n = 7,895 (81.4%) patients included in the analysis.

**Table 4.** The association between intraventricular conduction disorders and the incidence of sudden cardiac death among patients treated for acute coronary syndrome. Hazard ratio estimates (HRs) calculated with traditional Cox regression, which does not take competing events due to death due to other reasons into account.

| Hazard ratio for sudden cardiac death |  | HR (95% CI) | P value |
| --- | --- | --- | --- |
| Unadjusted | QRS 110–119 ms | 2.28 (1.76–2.95) | < 0.001 |
|  | RBBB | 1.61 (1.09–2.39) | 0.017 |
|  | LBBB | 1.91 (1.13–3.28) | 0.017 |
|  | NIVCD | 6.40 (4.54–9.01) | < 0.001 |
| Age- and sex-adjusted | QRS 110–119 ms | 2.11 (1.63–2.74) | < 0.001 |
|  | RBBB | 1.34 (0.90–1.99) | 0.15 |
|  | LBBB | 1.71 (1.00–2.92) | 0.052 |
|  | NIVCD | 5.31 (3.75–7.52) | < 0.001 |
| Age-, sex-, and risk-factor-adjusted* | QRS 110–119 ms | 1.98 (1.51–2.58) | < 0.001 |
|  | RBBB | 1.21 (0.81–1.80) | 0.36 |
|  | LBBB | 1.33 (0.76–2.34) | 0.32 |
|  | NIVCD | 4.50 (3.16–6.40) | < 0.001 |
| Age-, sex-, and LVEF-adjusted** | QRS 110–119 ms | 1.84 (1.41–2.41) | < 0.001 |
|  | RBBB | 1.36 (0.91–2.04) | 0.14 |
|  | LBBB | 1.11 (0.63–1.94) | 0.73 |
|  | NIVCD | 3.65 (2.54–5.24) | < 0.001 |
| Age- and sex-adjusted (LVEF > 35%)* | QRS 110–119 ms | 1.83 (1.31–2.57) | < 0.001 |
|  | RBBB | 1.25 (0.77–2.02) | 0.38 |
|  | LBBB | 1.01 (0.41–2.45) | 0.99 |
|  | NIVCD | 4.62 (2.87–7.43) | < 0.001 |

\*, \*\*, and \*\*\*: same as in Table 3.

**Table 5.** Effect of the onset of conduction disorders (old vs new/presumabely new) on the association with the incidence of sudden cardiac death among patients treated for acute coronary syndrome. Hazard ratio estimates (HRs) correspond to subdistribution hazard estimates analyzed by both Fine-Gray subdistribution hazard modeling and Cox regression.

| Hazard ratio for sudden cardiac death |  | HR (95% CI) | P value |
| --- | --- | --- | --- |
| Unadjusted (SDH) | QRS 110–119 ms | 2.11 (1.63–2.73) | < 0.001 |
|  | RBBB (old) | 0.83 (0.43–1.60) | 0.57 |
|  | RBBB (new) | 1.78 (1.11–2.85) | 0.016 |
|  | LBBB (old) | 1.18 (1.11–2.85) | 0.71 |
|  | LBBB (new) | 2.01 (1.03–3.92) | 0.040 |
|  | NIVCD (old) | 4.09 (2.50–6.71) | < 0.001 |
|  | NIVCD (new) | 4.02 (2.54–6.36) | < 0.001 |
| Age- and sex-adjusted (SDH) | QRS 110–119 ms | 1.94 (1.50–2.53) | < 0.001 |
|  | RBBB (old) | 0.75 (0.38–1.47) | 0.40 |
|  | RBBB (new) | 1.65 (1.031–2.65) | 0.037 |
|  | LBBB (old) | 1.14 (0.47–2.80) | 0.77 |
|  | LBBB (new) | 2.02 (1.03–3.96) | 0.040 |
|  | NIVCD (old) | 3.69 (2.23–6.11) | < 0.001 |
|  | NIVCD (new) | 3.68 (2.31–5.84) | < 0.001 |
| Unadjusted (cox Regression) | QRS 110–119 ms | 2.28 (1.76–2.95) | < 0.001 |
|  | RBBB (old) | 1.08 (0.56–2.10) | 0.81 |
|  | RBBB (new) | 2.13 (1.33–3.43) | 0.002 |
|  | LBBB (old) | 1.45 (0.60–3.50) | 0.41 |
|  | LBBB (new) | 2.35 (1.21–4.55) | 0.012 |
|  | NIVCD (old) | 6.53 (4.00–10.65) | < 0.001 |
|  | NIVCD (new) | 6.28 (3.99–9.88) | < 0.001 |
| Age- and sex-adjusted (cox Regression) | QRS 110–119 ms | 2.11(1.62–2.74) | < 0.001 |
|  | RBBB (old) | 0.86 (0.44–1.67) | 0.65 |
|  | RBBB (new) | 1.85 (1.15–2.98) | 0.011 |
|  | LBBB (old) | 1.22 (0.52–2.96) | 0.66 |
|  | LBBB (new) | 2.18 (1.21–4.23) | 0.022 |
|  | NIVCD (old) | 5.18 (5.18–3.16) | < 0.001 |
|  | NIVCD (new) | 5.38 (3.41–8.48) | < 0.001 |

In a sensitivity analysis, when the analysis was limited to patients without ICDs, the SCD risk was similarly associated with NIVCD and a prolonged QRS duration of 110–119 ms (Supplementary Table 3). Only prolonged QRS duration was not associated with SCD in patients with no ICD with a LVEF of > 35%. Additionally, the results remained essentially unchanged when SCA was used as an endpoint (Supplementary Table 4).

### Onset of intraventricular conduction disorders and SCD events during follow-up

The time of onset of intraventricular conduction disorders (pre-existing before ACS or presumably new) and their relation to SCD were investigated in a separate analysis (Table 4). According to the survival analysis of these subgroups, both old and presumably new NIVCDs were associated with SCD. As for LBBB and RBBB, a significant association was observed only for new LBBB and new RBBB.

## Discussion

In the present large study with consecutive ACS patients, all of whom were undergoing modern invasive evaluation for ACS, we found 1) NIVCD, defined as a QRS duration of ≥ 120 ms without meeting the criteria for LBBB or RBBB, to be a powerful predictor of SCD independently of clinical risk factors and across the subcategories of LVEF. We also found 2) a QRS duration of 110–119 ms to similarly predict SCD, and that 3) patients with NIVCD and a QRS duration of 110–119 ms together accounted for 23% of all SCDs in the study. Furthermore, 4) both old and presumably new NIVCD predicted SCD, while only presumably new RBBB or LBBB associated with arrhythmic death.

### Conduction disorders and SCD

The fact that NIVCD is related to strikingly high rates of SCDs independently of baseline LVEF in patients who are not eligible for a primary-preventive ICD suggests that there is novel potential to improve preventive srtategies for arrhythmic deaths in ACS patients. We observed that SCD events were significantly overrepresented in NIVCD and the risk of SCD was over three-fold, even in the subgroups with a LVEF of > 35 % (i.e., in patients not meeting the conventional criterion for a primary-preventive ICD device) and in those who never received an ICD device for any indication.

Prior investigators have associated NIVCD with arrhythmic death in high-risk patients with ventricular irritability and LVEF < 40 % (31). In a smaller case-control study of patients with documented coronary artery disease, NIVCD was associated with SCD (32). Aro et al. associated NIVCD with arrhythmic death in a general population cohort enrolled from 1966 to 1972 (13). Furthermore, in a primary care cohort with 326,227 individuals, LBBB and NIVCD were identified as the strongest ECG predictors of out-of-hospital cardiac arrest (19). Although the two previously mentioned population-based studies were unable to control for all relevant SCD risk factors, such as left ventricular function, they corroborate our observations.

Previous studies and pacing guidelines have defined non-specific IVCD as either a QRS duration of ≥ 110 ms (13,31,33) or a QRS duration of ≥120 ms (19,32,34) without meeting the criteria for LBBB or RBBB. Thus, we decided to include QRS duration of 110–119 ms as a covariate in our study. We found both definitions of NIVCD to predict SCD, albeit the risk of arrhythmic death was significantly more pronounced in those with a QRS duration of ≥ 120 ms.

New-onset LBBB or RBBB in the setting of ACS has previously been found to predict cardiovascular death (35–37). These events include deaths due to heart failure and sudden arrhythmic deaths, but also deaths due to other cardiovascular complications of ACS. The risk of SCD in those with new-onset NIVCD has not been studied before. While we found both new LBBB and RBBB to predict SCD, the highest risk of arrhythmic death was, again, observed in those with NIVCD. However, whether the NIVCD pattern was a result of a recent acute myocardial infarction or past underlying myocardial cardiopathy bears no additional significance for risk stratification, as both old and new NIVCD were robust predictors of SCD in our study.

### SCD prevention with an ICD

Although new ICD device implantations during the study follow-up were more prevalent in those with LBBB and NIVCD, the findings from the present study confirm the need to identify other SCD risk predictors besides severe left ventricular dysfunction in post-infarction patients.

The current clinical approach to the primary prevention of SCD relies mainly on the provision of an ICD to patients with severely reduced left ventricular function (LVEF < 35%) (38). The major problem with this approach is that enrolment for the studies on which this approach is based was carried out almost three decades ago (39,40), while both pharmacological and interventional treatments for ACS and heart failure have improved substantially since then (38). Additionally, the majority (∼ 70%) of SCD victims do not have a severely reduced LVEF (12,41). In other words, prior randomized trials show that a reduced LVEF is associated with an increased SCD risk and that ICD therapy improves survival, but they do not establish LVEF as the optimal risk stratification variable. Therefore, there is an obvious need for better risk stratification tools for SCD, especially in patients with a history of myocardial infarction (41,42).

The 12-lead electrocardiogram (ECG) is an attractive tool for SCD risk evaluation. While NIVCD, which is an easily recognized ECG finding, was found to be highly predictive of SCD in our study, the matter of whether its presence could trigger the implantation of an ICD device in the primary prevention of SCD in post-ACS patients warrants further study and a randomized trial. Unfortunately, it appears that a randomized ICD (PROFID-Preserved) trial of post-MI patients with a preserved LVEF of > 35% but a high risk of SCD was recently withdrawn, as no prior available risk predictors with an acceptable predictive performance for the discrimination of high-risk patients were found (NCT04540289).

### Strenghts and limitations

Our study provides an update on the prognosis of bundle branch blocks and, most importantly, solidifies the significance of NIVCD as a predictor of SCD. The present study population comprises all consecutive patients treated in a limited geographical area during a 14-year period.

Consequently, this population can be considered to be an unselected and unbiased representation of patients invasively diagnosed with ACS. Furthermore, given the large overall sample size resulting in the largest study to date with post-ACS patients with NIVCD, the study has sufficient power to detect significant associations despite the small relative size of the each patient group. The determination of SCD was based on an in-depth review of medical records and death certificates. We excluded sudden death cases that were not of cardiac causes, as well as those which were not unexpected, such as in the case of prolonged cardiac symptoms or a vague event description. While our study was retrospective, the definition of ECG features was decided *a priori,* and the adjudication of conduction disorders observed in ECG recodrings in our study was conducted by investigators blinded to the outcome. Additionally, we also used a subdistribution hazard model, which is more accurate than traditional Cox regression in describing the real risk of SCD (30).

As for limitations, the fact that non-invasively diagnosed patients were not included in this study means that the prognosis of these patients cannot be assessed. However, this exclusion was made because the diagnosis of ACS in patients not undergoing an invasive evaluation is uncertain due to possible confounding issues caused by type II myocardial infarctions and because, with these patients, the reason for adopting a non-invasive strategy was usually based on a poor overall prognosis (20). Additionally, despite adequately controlling for clinical risk factors, the presence of additional residual confounding cannot be excluded. Finally, our results show the importance of distinguishing NIVCD, but we do acknowledge that, in some cases, differentiation between LBBB and NIVCD is challenging in a real-life setting. To mitigate this, we used the modern definition of LBBB introduced by Strauss et al., which is stricter in that many ECG-reported cases of LBBB have a wide QRS complex due to a combination of LAHB and LVH (i.e., NIVCD) (26).

## Conclusion

We found NIVCD (QRS ≥ 120 ms) to be a robust predictor of SCD without meaningful elevations in competing deaths across the LVEF spectrum. A more permissive threshold (QRS ≥ 110 ms according to current guidelines (27)) for NIVCD resulted in a similar but slightly lower risk of SCD but accounted for 23% of all SCD cases in the study. Further research and a randomized trial are needed on ICD as primary prevention of SCD in post-ACS patients with NIVCD. Furthermore, our results support the current guidelines describing new-onset LBBB and RBBB as a high-risk feature in ACS patients.

## Data Availability

Data available in anonymous form pending the approval of the study steering committee.

---

**Supplementary Table 1:** The classification of NIVCDs by the 12SL Marquette system compared to clinical evaluation.

|  | GE Marquette |  |  |  |
| --- | --- | --- | --- | --- |
| Clinical | RBBB<br>n = 537 | LBBB<br>n = 281 | NIVCD<br>n = 172 | QRS ≥ 120 ms<br>n = 251 |
| QRS < 120 ms, n = 7,847 | 14 (2.6%) | 3 (1.1%) | 24 (14%) | 187 (74.5%) |
| RBBB n = 530 | 523 (97.4%) | 0 | 5 (2.9%) | 2 (0.8%) |
| LBBB n = 227 | 0 | 217 (77.2%) | 8 (4.7%) | 2 (0.8%) |
| NIVCD n = 256 | 0 | 61 (21.7%) | 135 (78.5%) | 60 (23.9%) |

clinical evaluation
GE Marquette's analysis was the same as the clinical analysis in iRBBB (n = 15) and iLBBB (n = 93). 93/293 (31.7%) LAFB patients also had RBBB. 11/16 (68.8%) LAFB patients also had RBBB. 15 LAFBs and 1 LPFB were classified as QRS ≥ 120 ms, but after clinical evaluation, only 3 LAFBs were reclassified as NIVCD and the other LAFBs/LPFBs as QRS < 120 ms. There were no reclassifications regarding LAFB/LPFB classified by GE Marquette as QRS < 120m.

**Supplementary Table 2.** GE Marquette’s diagnostic tests, assuming that GE Marquette does not misanalyse the QRS duration incorrectly as under 120.

| GE Marquette | Sensitivity | Specificity | Positive predictive value | Negative predictive value |
| --- | --- | --- | --- | --- |
| RBBB | 98.7% | 99.9% | 97.4% | 99.9% |
| LBBB | 95.6% | 99.3% | 77.2% | 99.9% |
| NIVCD | 52.7% | 99.6% | 78.5% | 98.7% |

**Supplementary Table 3.** The association between intraventricular conduction disorders and the incidence of sudden cardiac death among patients treated for acute coronary syndrome who never received an intracardiac defibrillation device (n = 9,385). Hazard ratio estimates (HRs) correspond to subdistribution hazard estimates analyzed by Fine-Gray subdistribution hazard modeling, accounting for competing events due to deaths due to other causes.

| Hazard ratio for sudden cardiac death |  | HR (95% CI) | P value |
| --- | --- | --- | --- |
| Unadjusted | QRS 110–119 ms (n = 795, 94.6%) | 1.93 (1.43–2.60) | < 0.001 |
|  | RBBB (n = 504, 95.1%) | 1.11 (0.70–1.77) | 0.65 |
|  | LBBB (n = 197, 86.8%) | 0.91 (0.40–2.05) | 0.82 |
|  | NIVCD (n = 223, 87.1%) | 3.92 (2.63–5.85) | < 0.001 |
| Age- and sex-adjusted | QRS 110–119 ms | 1.79 (1.33–2.42) | < 0.001 |
|  | RBBB | 0.98 (0.61–1.57) | 0.92 |
|  | LBBB | 0.83 (0.37–1.89) | 0.66 |
|  | NIVCD | 3.37 (2.23–5.07) | < 0.001 |
| Age-, sex-, and risk-factor-adjusted* | QRS 110–119 ms | 1.61 (1.18–2.20) | 0.003 |
|  | RBBB | 0.89 (0.55–1.43) | 0.63 |
|  | LBBB | 0.63 (0.26–1.53) | 0.31 |
|  | NIVCD | 2.74 (1.79–4.20) | < 0.001 |
| Age-, sex-, and LVEF-adjusted** | QRS 110–119 ms | 1.71 (1.25–2.32) | < 0.001 |
|  | RBBB | 0.99 (0.61–1.61) | 0.96 |
|  | LBBB | 0.60 (0.24–1.49) | 0.27 |
|  | NIVCD | 2.71 (1.75–4.18) | < 0.001 |
| Age- and sex-adjusted (LVEF > 35%)*** | QRS 110–119 ms | 1.38 (0.93–2.05) | 0.11 |
|  | RBBB | 0.82 (0.45–1.47) | 0.50 |
|  | LBBB | 0.85 (0.31–2.29) | 0.80 |
|  | NIVCD | 3.03 (1.74–5.30) | < 0.001 |

\*Model adjusted with age, sex, prevalent valvular heart disease, prevalent diabetes, previous coronary artery bypass grafting, serum creatinine value, prevalent peripheral artery disease, history of atrial fibrillation, and history of stroke. n= 9,272 (98.8% of patients with no ICD) patients included in the analysis.
\*\*n = 8,665 (92.3%) patients included in the analysis.
\*\*\*n = 7,721 (82.3%) patients included in the analysis.

**Supplementary Table 4.**
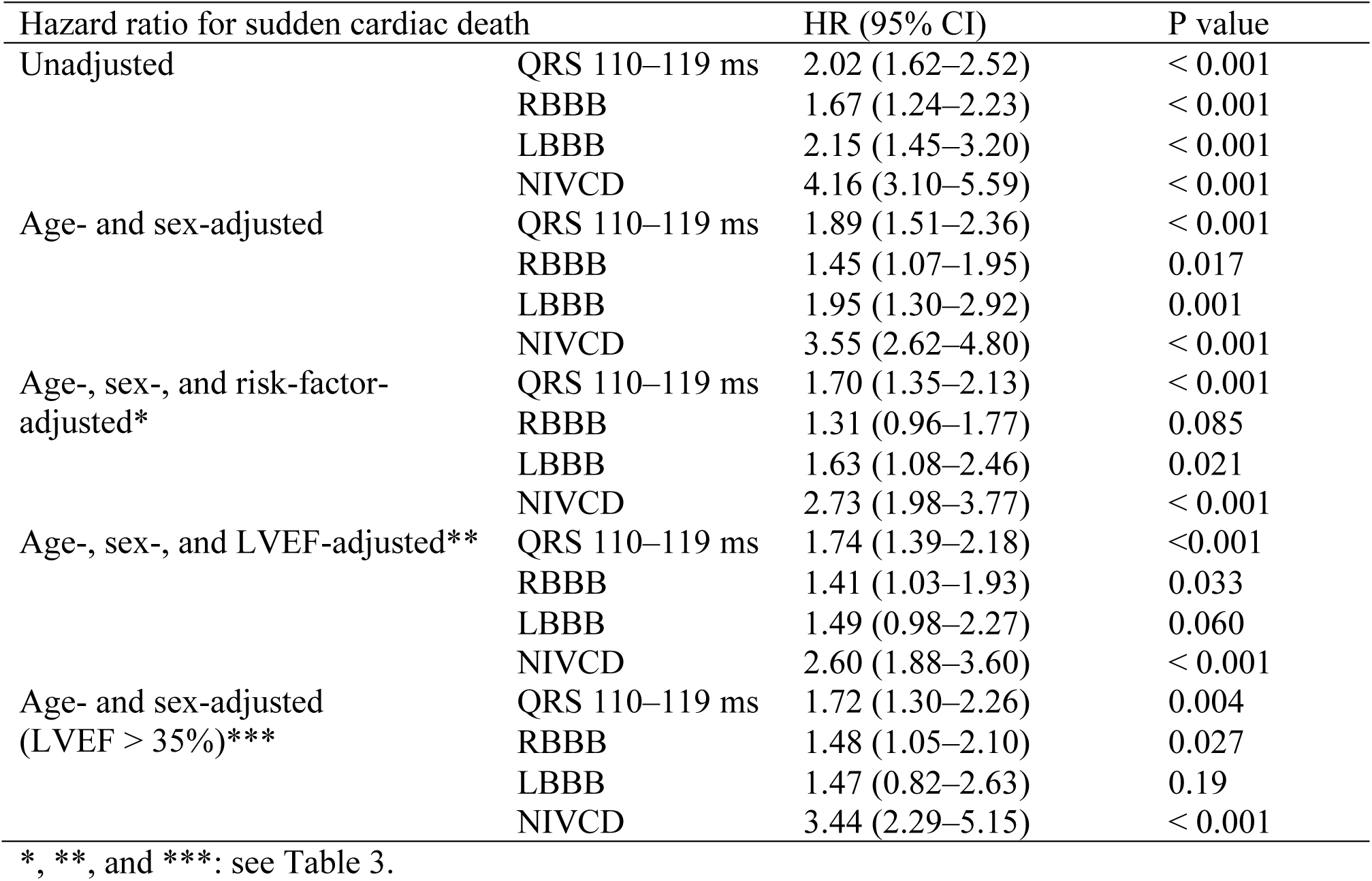
The association between intraventricular conduction disorders and the incidence of sudden cardiac arrest among patients treated for acute coronary syndrome. Hazard ratio estimates (HRs) correspond to subdistribution hazard estimates analyzed by Fine-Gray subdistribution hazard modeling.

